# A satellite-based spatio-temporal machine learning model to reconstruct daily PM_2.5_ concentrations across Great Britain

**DOI:** 10.1101/2020.07.19.20157396

**Authors:** Rochelle Schneider dos Santos, Ana M. Vicedo-Cabrera, Francesco Sera, Pierre Masselot, Massimo Stafoggia, Kees de Hoogh, Itai Kloog, Stefan Reis, Massimo Vieno, Antonio Gasparrini

## Abstract

Epidemiological studies on health effects of air pollution usually rely on measurements from fixed ground monitors, which provide limited spatio-temporal coverage. Data from satellites, reanalysis and chemical transport models offer additional information used to reconstruct pollution concentrations at high spatio-temporal resolution. The aim of this study is to develop a multi-stage satellite-based machine learning model to estimate daily fine particulate matter (PM_2.5_) levels across Great Britain during 2008-2018. This high-resolution model consists of random forest (RF) algorithms applied in four stages. Stage-1 augments monitor-PM_2.5_ series using co-located PM_10_ measures. Stage-2 imputes missing satellite aerosol optical depth observations using atmospheric reanalysis models. Stage-3 integrates the output from previous stages with spatial and spatiotemporal variables to build a prediction model for PM_2.5_. Stage-4 applies Stage-3 models to estimate daily PM_2.5_ concentrations over a 1 km grid. The RF architecture performed well in all stages, with results from Stage-3 showing an average cross-validated R^2^ of 0.767 and minimal bias. The model performed better over the temporal scale when compared to the spatial component, but both presented good accuracy with an R^2^ of 0.795 and 0.658, respectively. The high spatio-temporal resolution and relatively high precision allows this dataset (approximately 950 million points) to be used in epidemiological analyses to assess health risks associated with both short- and long-term exposures to PM_2.5_.

## 1. Introduction

The World Health Organization estimates in 7 million the global deaths associated with air pollution (both outdoor and household) every year, emphasising that exposure to particulate matter (PM) is among the greatest cause of concern [WHO,2020]. Fine particles with aerodynamic diameter smaller than 2.5 μm (PM_2.5_) can penetrate into the human circulatory system through the lungs and provoke multiple adverse health outcomes, including mortality [Liu et al., 2019] hospital admissions [Basagaña, et al., 2015], lung dysfunction [Raaschou-Nielsen et al., 2016], cardiovascular diseases [Lavigne, et al., 2019b], and allergic reactions [Lavigne et al., 2019a]. Usually, epidemiological studies collect air quality (AQ) data from ground monitors to quantify both short-term and long-term PM_2.5_ exposures associated with acute and chronic health effects, respectively. The limitation in this health assessment approach is the lack of continuous temporal records of PM_2.5_ and the limited spatial distribution of the monitors. Great Britain is an example of countries with very limited spatio-temporal coverage of PM_2.5_, whereby the monitoring network is densely located only in major cities and widespread measurements of PM_2.5_ only started from 2010.

Remote sensing observations of aerosol optical depth (AOD) obtained from satellites, which measures how much direct sunlight has been scattered and absorbed by aerosols particles suspended in the atmosphere, has recently been proposed as an alternative to measure PM variability for epidemiological purposes [NASA Earth Observations, 2020]. However, while offering the advantage of global coverage and relatively high spatio-temporal resolution, the use of AOD for PM_2.5_ exposure assessments present limitations, for instance the fact that it represents the total atmospheric column concentration of aerosol rather than surface values [van Donkelaar, 2010]. Unsurprisingly, early studies based only on satellite-AOD achieved very low performances in predicting PM_2.5_ [Koelemeijer et al., 2006]. Recent studies have proposed more sophisticated approaches, combining AOD measures with information from other satellite products, reanalysis data, chemical transport models, and geospatial features to improve the prediction of PM_2.5_. Such studies used various analytical methods, including multiple linear regression [Gupta and Christopher, 2009], land-use regression [Beckerman et al., 2013; Vienneau et al., 2010], and mixed effect models [de Hoogh et al., 2018; Kloog et al., 2011; Kloog et al., 2015; Lee et al., 2011; Staffogia et al., 2016]. The last development in this research area is represented by the application of machine learning (ML) algorithms, including various architectures such as random forests [Chen et al., 2018; Di et al., 2019; Staffogia et al., 2019; Yazdi, et al., 2020], neural network [Di et al., 2019; Yazdi, et al., 2020], and gradient boosting [Chen et al., 2019; Di et al., 2019; Just et al., 2018; Zan et al., 2017]. These have demonstrated higher performances, linked with the ability to model any kind of predictor(s)-response association and to deal better with potentially complex relationship between PM_2.5_, spatial, and spatio-temporal predictors [Di, et al., 2019; Polley et al., 2010].

The aim of this study is to develop and apply a multi-stage satellite-based ML model to estimate daily concentrations of PM_2.5_ over a 1 km grid across Great Britain in the period 2008-2018. The analysis is based on a dataset with synchronised information from various data sources, such as several remote sensing satellite products, multiple climate and atmospheric reanalysis databases, chemical transport models, and spatial and spatio-temporal variables. The model is assessed through measures of predictive performance, error, and bias, obtained through cross-validation.

## 2. Materials and Methods

### 2.1. Study area and period

Great Britain is an island with an extension of 229,462 km^2^ surrounded by the Atlantic Ocean, Irish Sea, North Sea, and English Channel. It comprises the countries of England, Scotland, and Wales with a total population in 2018 of almost 65 million [ONS, 2020]. According to the Köppen climate classification, the United Kingdom (Great Britain and Northern Ireland) is defined as having a warm temperate climate, fully humid with mostly warm summer (cold summer for some parts of Scotland and England) [Kottel et al., 2006]. The study area included 234.429 1 km grid cells (containing a unique identification code, cell-ID) from the original 1 km Great Britain National Grid Squares [Digimap, 2020] for a period between 1^st^ January 2008 and 31^st^ December 2018.

### 2.2. PM_2.5_ and PM_10_ observed data

Daily PM_10_ and PM_2.5_ (µg/m^3^) measurements in the study period were obtained from five monitoring network sources through the R package *openair* [Openair, 2020]: Automatic Urban andRural Network (AURN), Air Quality England (AQE), King College London (KCL), Scotland Air Quality Network (SAQN), and Wales Air Quality Network (WAQN). When monitors from different sources showed exactly the same temporal distributions *(i.e*., correlation equal to 1) and were located at approximately the same coordinates, only the AURN monitor was kept. Monitors with less than 18 hours of PM_2.5_ records per day as well as less than 30 days by year were removed. The final set includes 581 and 183 monitors measuring PM_10_ and PM_2.5_ along the study period, respectively. Figure 1 shows the locations of these monitors across Great Britain, illustrating how the network coverage is densely located in major cities, leaving several small cities and rural areas with only a few or none AQ records. Each monitor was indexed using the cell-ID of the 1 km grid cell that contains it.

**Figure 1:**
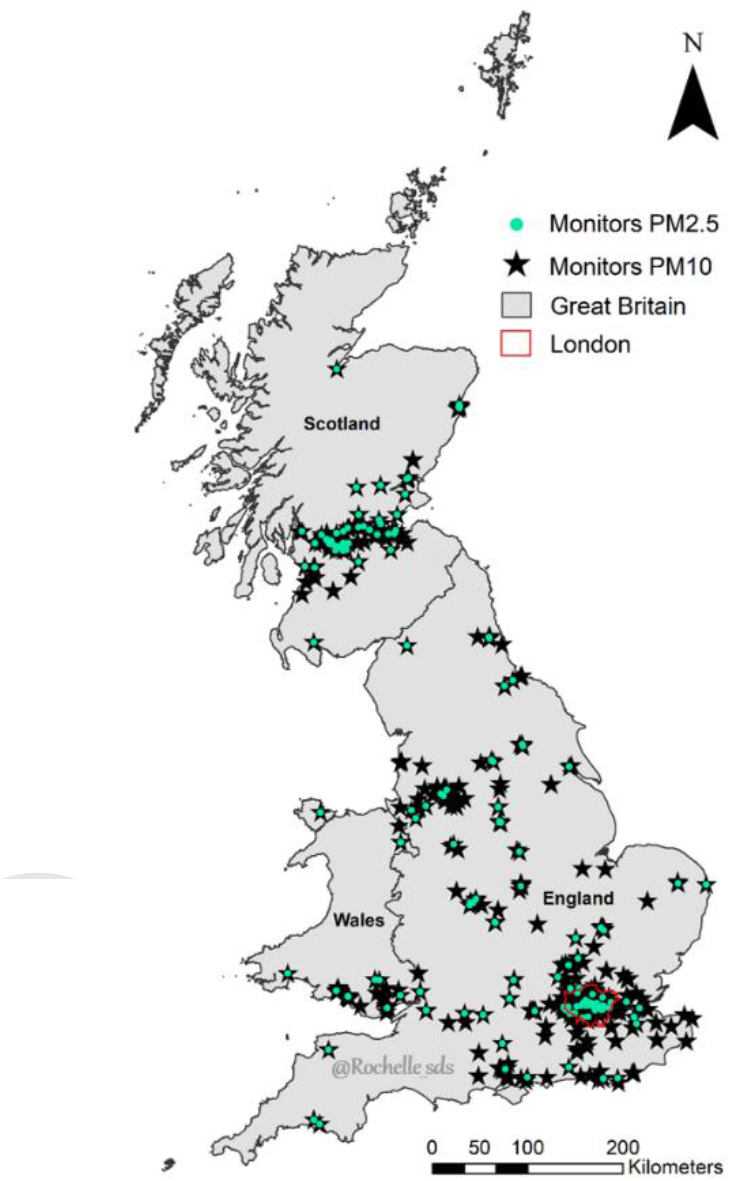
Spatial distribution of 581 PM_10_ (black star) and 183 PM_2.5_ (turquoise dots) monitors across Great Britain during the study period.

### 2.3. Spatially-lagged and nearest monitor PM_2.5_ variables

Four spatio-temporal variables were generated from the monitor series of PM_2.5_ to represent spatially-lagged annual average concentrations. Monitor types were grouped into two classes (background (urban, suburban, and rural) and hotspots (traffic and industrial) to compute the annual averages values of nearby monitors by class using inverse-distance weighted leave-one-out cross-validated (IDW-LOOCV) approach. Two different weights were applied, namely the inverse distance and the inverse squared distance (in km). The former assigns relatively more weight to distant monitors, and therefore can represent a *regional* background, while the latter captures *local* differences. Therefore, these four IDW-LOOCV variables were named as follow: (i) Spatially-lagged hotspot-PM_2.5_ regional, (ii) Spatially-lagged background-PM_2.5_ regional, (iii) Spatially-lagged hotspot-PM_2.5_ local, and (iv) Spatially-lagged background-PM_2.5_ local. To improve the model performance in the spatial domain, two additional spatial variables were generated based on the closest Euclidean distance for each monitor class, named as: (i) Nearest hotspot monitor distance and (ii) Nearest background monitor distance. These six variables were used as additional predictors to capture the heterogeneity across monitors and exploit their spatial autocorrelation, and thus help the model to better categorise differences in the spatial patterns of measured-PM_2.5_ series.

### 2.4. AOD data: satellite and atmospheric reanalysis models

Daily satellite-AOD was obtained from the Collection 6 Level-2 gridded product (MCD19A2). These data are generated at a 1 km grid through the Multi-angle Implementation of Atmospheric Correction (MAIAC) algorithm using data from Moderate Resolution Imaging Spectroradiometer (MODIS) sensor on board both Terra and Aqua earth observation satellites [Lyapustin and Wang, 2018]. Four layers from MCD19A2 product were extracted: (i) AOD Blue band (0.47μm), (ii) AOD Green band (0.55μm), (iii) AOD uncertainty (i.e. level of uncertainty based on blue-band surface brightness [reflectance]), (iv) AOD_QA (quality assurance flags to retrieve only the best quality AOD). These layers are generated for each passing time of Terra and Aqua satellites over the area of study and combined by day. The layers AOD-0.47μm and AOD-0.55μm were used as the outcome variables, after their values were filtered using AOD uncertainty and AOD_QA layers to guarantee high product quality.

This calibration process, together with pixels covered by clouds, removed a large sample of AOD grid cells over Great Britain. To fill the satellite-AOD gaps, modelled-AOD total column was used from Copernicus Atmosphere Monitoring Service (CAMS) reanalysis provided by the European Centre for Medium-Range Weather Forecasts (ECMWF) [Bozzo, et al., 2017]. CAMS reanalysis provides every 3-hourly modelled-AOD at five different wavelengths (0.47µm, 0.55µm, 0.67µm, 0.87μm, and 1.24µm) with a spatial resolution of approximately 80 km but the data was downloaded at 10 km, based on an interpolation performed through the ECMWF’s API request. The satellite-AOD and CAMS modelled-AOD were indexed to the closest 1 km grid cell from their pixel centroid.

### 2.5. Other spatio-temporal predictors

#### 2.5.1. Modelled PM_2.5_ from chemical transport models

Atmospheric chemistry transport models (ACTMs) incorporate anthropogenic and natural sources of emission, land use and meteorological conditions to simulate the atmospheric compositions and deposition of various air pollutants (trace gases and particles). Based on the European Modelling and Evaluation Programme (EMEP) ACTM, EMEP4UK, has been developed to represent the UK hourly atmospheric composition at a spatial resolution of 5 km [EMEP4UK, 2020]. The description of EMEP4UK model framework and setup can be found elsewhere [Vieno et al., 2010]. Daily EMEP4UK simulations of PM_2.5_ (µg/m^3^) concentrations at surface-level were included to represent ground-level contributions, in contrast to AOD products that refer to the total column of aerosols concentrations. Each EMEP4UK 5 km pixel was linked to the closest 1 km grid cell centroid.

#### 2.5.2. Meteorological variables from climate reanalysis models

Meteorological variables were retrieved from the ECMWF’s climate reanalysis models with the highest spatial resolution available during 2008-2018 and at two sub-day times (0:00 and 12:00). Sea pressure and the boundary layer height (BLH) were downloaded from the ERA 5 global reanalysis with a spatial resolution of approximately 30 km [ERA5, 2020]. Air temperature at 2m height and total precipitation were obtained from the ERA 5 Land global reanalysis with a spatial resolution of approximately 9 km [ERA5 Land, 2020]. Relative humidity, wind direction, and wind speed were downloaded from UERRA regional reanalysis at 5.5 km for the MESCAN-SURFEX system [UERRA, 2020]. All meteorological variables were indexed to the closest 1 km grid cell to their centroid.

#### 2.5.3. Normalized difference vegetation index

Monthly Normalized difference vegetation index (NDVI) layer is used to quantify vegetation presence and it ranges from -1 to 1. The NDVI 1 km grid was obtained from MOD13A3 Version 6 Level 3, Terra-MODIS product [Didan et al., 2015]. Each NDVI pixel was indexed to the closest 1 km grid cell to its pixel centroid and the NDVI values repeated for the days inside the corresponded month.

### 2.6. Spatial predictors

#### 2.6.1. Land variables and night-time light data from earth observation satellites

Three land predictors were collected from the Copernicus Land Monitoring Service (CLMS) [CLMS, 2020] database: elevation, land cover, and impervious surfaces. Elevation data was obtained from the 2011 European Digital Elevation Model (EU-DEM) version 1.1 with a spatial resolution of 25 m. The elevation values were obtained from the mean of all 25 m-pixels values located inside each 1 km grid cells. Land cover data was obtained from 2012 CORINE Land Cover (CLC) inventory. It was derived from high-resolution ortho-rectified satellites images that mapped all land elements at a spatial resolution ranging from 5 m to 60 m and aggregated into 100 m. Nine predictors were defined by grouping the original 44 CLC classes and each predictor represents its group proportion inside each 1 km Great Britain Grid cell. Imperviousness degree is a binary raster product at a spatial resolution of 100 m, where the value 0 represents natural land cover or water surface and value 1 represents entirely artificial surfaces (i.e. built-up areas). The amount of impervious surfaces was estimated by the proportion of artificial surfaces inside each 1 km Great Britain Grid cell.

Night-time lights data was provided by the visible Infrared Imaging Radiometer Suite (VIIRS) sensor aboard the Suomi-National Polar-orbiting Partnership (Suomi-NPP) satellite. The VIIRS Day/Night band collects cloud-free average radiance values at annual and monthly composites in spatial resolution of 750m [EOG, 2020]. The 2015 night-time lights annual mean was computed based on the weighted average of VIRRS pixels inside each 1 km Great Britain Grid cell.

#### 2.6.2. Population density

The resident population counts data from 2011 were collected from the Office for National Statistics (England and Wales) and the National Records of Scotland. The smallest geographic unit of the UK census is output areas (OA), with a total of 227.769 OAs polygons for Great Britain. The proportion of each OA’s area inside each 1 km Great Britain Grid cell was extracted to estimate the weighted average population by 1 km grid cell.

#### 2.6.3. Road density and distance

Road density and length predictors were derived from the Ordnance Survey (OS) [2020] Open Roads product, which offers a geospatial representation of Great Britain’s Road network. Three density predictors were defined by grouping the original eight OS road types, where each predictor was computed as the sum of all roads length inside each 1 km Great Britain Grid cell by group (highway, secondary, and local). Three distance predictors were defined by computing the inverse distance of each 1 km grid centroid from the closest road group (highway, secondary, and local).

#### 2.6.4. Inverse distance from airports and seashore

Information on location and size of airports was derived from the Civil Aviation Authority (CAA) [2020], which collects monthly statistics about air traffic movements for more than 60 UK Airports, including aviation activities for terminal passengers, commercial flights, and cargo tonnage. A total of 19 airports across Great Britain were selected based on a minimum of 1% for the annual percentage of passengers at the airport during 2015-2018. For each cell of the 1 km Great Britain grid the inverse distance from the closest airport was calculated.

The inverse distance from seashore was computed for each 1 km grid cell using the geographical information about the boundaries of England, Wales, and Scotland provided by the UK Data Service [2020].

### 2.7. Statistical methods

A four-stage model was developed to obtain daily PM_2.5_ concentrations for all 234.429 grid cells covering Great Britain. Each stage is described in detail below. Briefly, Stage-1 applies a random forest (RF) algorithm to predict PM_2.5_ concentrations in monitors with only PM_10_ records. Stage-2 uses RF models to impute missing satellite-AOD from Terra and Aqua satellites, using modelled-AOD from CAMS. Stage-3 combines the output from Stage-1 and Stage-2 with a list of spatial and spatio-temporal synchronised predictors to estimate PM_2.5_ concentrations at the locations of the monitors. Stage-4 uses the Stage-3 model to predict daily PM_2.5_ across the whole Great Britain.

#### 2.7.1. Random forest algorithm

RF is a supervised tree-based design ML algorithm which trains an ensemble of independent decision trees (or forest) in parallel. The final model accuracy is estimated by the performance average of all decision trees. There are two main advantages of the RF architecture (known as bagging ensemble method): first, it controls the bias-variance trade-off by feeding each tree model with two-thirds of the training set while one-third is left out for validation *(i.e*., out-of-bag (OOB)) [Schneider dos Santos, 2020]. When all decision trees receive the same amount and list of predictors, they become highly correlated, not solving the variance problem. Therefore, the second positive aspect from RF is that the number of predictors on each tree is less than the full list available and they are randomly selected. This approach changes the predictor placed on the top of the tree, generating different splits and internal nodes. The algorithm is then able to estimate an importance ranking by quantifying the amount of error decreased due to a split of a specific predictor [James et al., 2013].

The performance of the models in each stage was assessed using statistics based on OOB samples and then from a 10-fold cross-validation (CV) procedure based on monitors. In the latter, ten random groups of monitors were defined, and the complete outcome series in each group were predicted using a model fitted in the other nine. This procedure offers a measure of true predictive ability of the RF model in locations where no monitor is available. Measures of performance were generated by regressing the OOB or cross-validated predicted values on the observed series, and computing the R^2^, the root mean square error (RMSE), and intercept and slope of the prediction. These statistics were computed overall using the whole series, and then separated in spatial and temporal contributions. The former was computed using the averages of predicted and observed values across the series, and it offers a measure of performance in capturing long-term average PM_2.5_ concentrations. The latter was computed as daily deviations from the averages, and it quantifies the temporal variability explained by the model.

#### 2.7.2. Stage-1: increasing PM_2.5_, measurements using co-located PM_10_ monitors

The number of PM_2.5_ monitors across Great Britain at the beginning of the study period was relatively low, and even if the quantity has increased substantially after 2010, most of the monitors were mostly installed in major cities. Stage-1 aims to increase the number of spatio-temporal ground-level PM_2.5_ references using observations from co-located monitors measuring PM_10_, which are available in higher number of locations and better distributed across Great Britain. Specifically, in this stage we fitted a RF model in locations with both PM_2.5_ and PM_10_ measurements, using data from all years (2008-2018). The RF model for each year *y* is defined as:

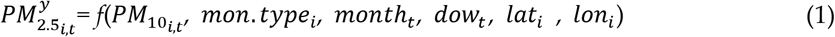

where: 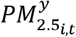 and 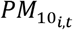 are the target variable and main predictor, respectively, measured in year *y* at monitor *i* on day t; *mon. type_i_* is a categorical variable classifying monitor *i* (traffic, industrial, urban, suburban, and rural); *month_t_* and *dow_t_* are categorical variables representing the months and day of the week of day t; and *lat_i_* and *lon_i_* define the coordinates of monitor *i*. The RF model was defined based on the best parameters setting. The optimised parameters were 500 decision trees as the RF ensemble (Ntree=500) and four variables randomly selected to be used on each tree (mtry=4). This model was eventually used to predict PM_2.5_ measurements in locations/days in which only PM_10_ was measured.

#### 2.7.3. Stage-2: imputing missing satellite-AOD from CAMS modelled-AOD

The percentage of missing satellite-AOD measurements in Great Britain ranged between 87% and 94% during 2008-2018 with the greatest portion during autumn and winter, near to the coast, and at the North of Scotland. Stage-2 imputes satellite-AOD missing for every day and 1 km grid based on an optimised RF model (Ntree=50 and mtry=20) and satellite-AOD wavelength (0.47µm and 0.55µm), separately in each year within the study period. These RF models were built for each year *y* as follows:

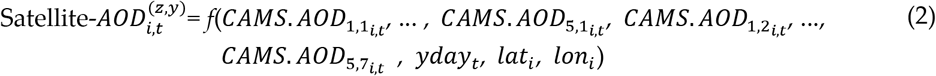

where: 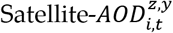 is the target variable representing satellite-AOD (wavelength *z*, year y) estimates at grid cell *i* on day t; CAMS.AOD is the main predictor representing CAMS modelled-AOD estimates at grid cell *i*, on day t, at five wavelengths (0.47µm, 0.55µm, 0.67µm, 0.865µm, and 1.24µm), and at seven sub-day times (3h, 6h, 9h, 12h, 15h, 18h, and 21h); *yday_t_* defines the sequence of days in a year from 1 to 365 (366, for leap years); *lat_i_* and *lon_i_* represent the coordinates of grid cell centroid *i*. This model was eventually used to predict missing satellite-AOD measurements.

#### 2.7.4. Stage-3: estimating PM_2.5_ concentrations using spatial and spatio-temporal variables

Stage-3 aims to build a predictive model for daily PM_2.5_ concentrations, using as target variable the combined set of PM_2.5_ directly measured from monitors or predicted from Stage-1, AOD measured from satellite instruments or predicted from Stage-2, together with all the spatially and spatio-temporally synchronized predictors described in the previous section. The RF models were fit separately by year *y*. The optimization process for this stage selected Ntree=500 and mtry=20 as the best set of parameter values. In this stage, PM_2.5_ concentrations were log-transformed to ensure the prediction of non-negative values. The Stage-3 model for each year *y* is defined as:

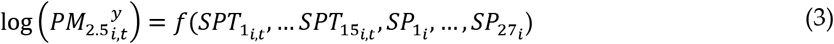

where: 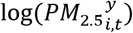 is the target variable representing the log-PM_2.5_ concentrations in year *y* at the monitor located in grid cell *i* on day *t*, while *SPT_it_* and *SP_i_* represent the spatio-temporal and spatial predictors, respectively.

Specifically, SPT_i,t_ included: surface-level log-PM_2.5_ values from the EMEP4UK model; Stage-2 AOD (at 0.47µm, 0.55µm); meteorological variables (air temperature, sea pressure, relative humidity, total precipitation, wind speed and direction); BLH (at 12:00 and 24:00 hours); monthly-averaged NDVI; and day of the year, month, and week. SPi included: spatially-lagged regional and local log-PM_2.5_ average concentrations from background and hotspot groups; nearest background and hotspot monitor distance; land variables (elevation, land cover in 9 groups, and % of impervious surface); night-time light; road density and inverse distance (each for highway, secondary, and local); population density; and inverse distance from closest airport and seashore.

#### 2.7.5. Stage-4: reconstructing PM_2.5_ time-series at 1 km grid

Using the RF models developed by year in Stage-3, daily PM_2.5_ concentrations for each 1 km grid cell were reconstructed across Great Britain for the whole study period (2008-2018).

## 3. Results

### 3.1. Stage-1 results

Not all monitors in each network provided the full series of daily PM concentrations during 2008-2018. However, PM_10_ was more often available, especially in the years 2008-2009, when there were less than 80 PM_2.5_ monitors across Great Britain. The imputation process in Stage-1 enabled the expansion from 46 to 269 in 2008 and from 65 to 278 in 2009. Table 1 shows the Stage-1 model performance (*i.e*., a separate RF model for each year) reported by two CV methods: (i) OOB - the original RF CV algorithm, which presented better accuracy (R^2^ average of 0.932), and (ii) 10-Fold CV - a targeted sampling, which leaves-out monitors with the full set of observations (R^2^ average of 0.855). As expected, the 10-Fold CV approach resulted in a slightly lower predictive accuracy although still showing a good performance. There is some indication of bias in the 10-Fold CV, with a slope lower than the expected value of 1 and a slightly negative intercept. The results for 10-Fold CV spatial and temporal domains are in Table A1 in Appendix A.

**Table 1.** Predicted-PM_2.5_ concentrations obtained from Stage-1 RF models were regressed against measured-PM_2.5_ concentrations in a linear regression model. The performance was evaluated using two CV methods (OOB and 10-Fold) together with RMSE (measure of the model error, µg/m^3^), intercept (µg/m^3^), and slope (µg/m^3^).

| Stage-1 |  |  |  |  |  |  |  |  |
| --- | --- | --- | --- | --- | --- | --- | --- | --- |
|  | OOB-CV |  |  |  | 10-Fold CV |  |  |  |
|  | R2 | RMSE | Inter. | Slope | R2 | RMSE | Inter. | Slope |
| 2008 | 0.918 | 1.196 | -0.417 | 1.033 | 0.707 | 4.954 | 0.803 | 0.886 |
| 2009 | 0.921 | 1.110 | -0.390 | 1.030 | 0.791 | 3.996 | 0.410 | 0.937 |
| 2010 | 0.919 | 1.122 | -0.401 | 1.029 | 0.843 | 3.496 | 0.043 | 0.983 |
| 2011 | 0.949 | 1.124 | -0.266 | 1.019 | 0.902 | 3.439 | -0.087 | 0.997 |
| 2012 | 0.942 | 1.058 | -0.274 | 1.021 | 0.889 | 3.218 | -0.035 | 0.986 |
| 2013 | 0.929 | 1.087 | -0.368 | 1.028 | 0.847 | 3.584 | 0.218 | 0.972 |
| 2014 | 0.944 | 0.963 | -0.267 | 1.022 | 0.891 | 3.003 | -0.007 | 0.995 |
| 2015 | 0.933 | 0.865 | -0.265 | 1.026 | 0.871 | 2.662 | -0.003 | 0.983 |
| 2016 | 0.935 | 0.896 | -0.251 | 1.025 | 0.885 | 2.654 | -0.050 | 0.996 |
| 2017 | 0.939 | 0.828 | -0.196 | 1.022 | 0.895 | 2.430 | 0.010 | 0.985 |
| 2018 | 0.928 | 0.791 | -0.248 | 1.028 | 0.886 | 2.235 | -0.019 | 0.993 |
| **Mean** | 0.932 | 1.003 | -0.304 | 1.026 | 0.855 | 3.243 | 0.117 | 0.974 |

### 3.2. Stage-2 results

The Stage-2 procedure is illustrated in Figure 2, showing missing satellite-AOD, imputed through a combination of multiple modelled-AOD wavelengths and sub-day times. Table 2 shows the performance of Stage-2 models validated using the OOB CV method. The results presented consistently high R^2^ (ranging from 0.963 to 0.989), low RMSE (varying between 0.006 to 0.011), and almost no bias (intercept zero and slope close to 1 in almost all years and wavelengths).

**Figure 2:**
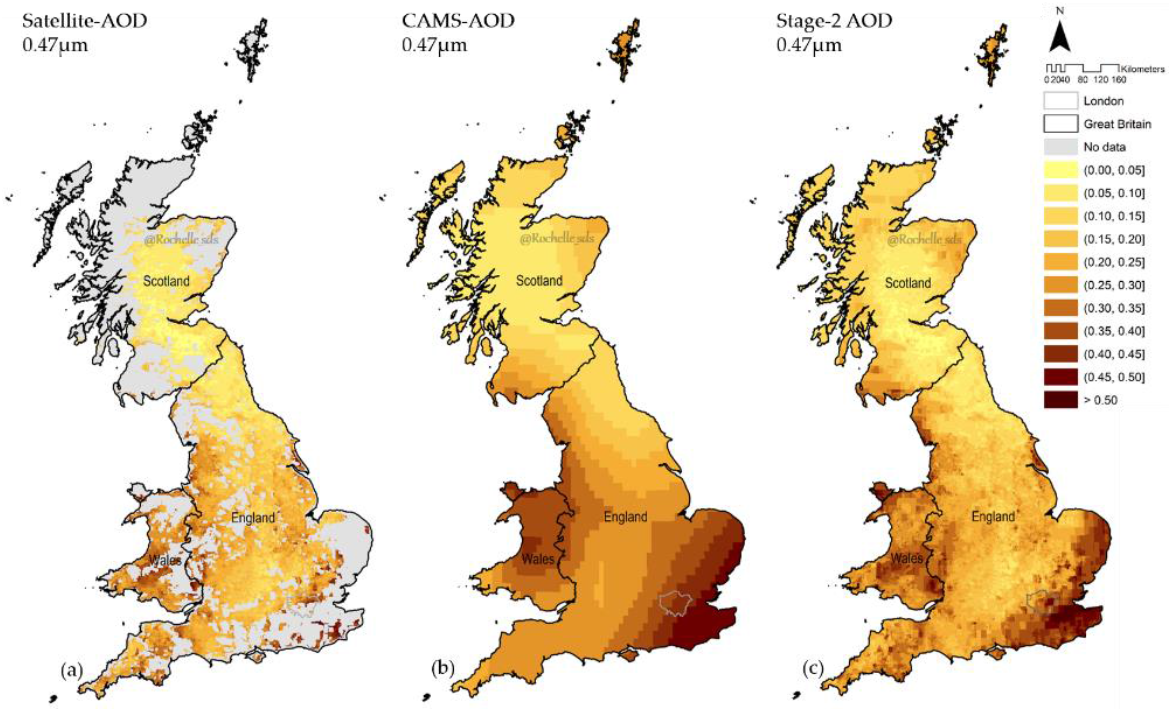
Satellite-AOD 0.47µm values are represented in Figure 2a (mean of all Terra- and Aqua-MODIS passing times). The missing values in grey were imputed through a combination of multiple modelled-AOD wavelengths and sub-day times, represented in Figure 2b by modelled-AOD 0.47µm at 12:00. Stage-2 output is shown in Figure 2c, illustrating the full coverage of AOD 0.47µm based on the combination of measurements and estimations across Great Britain. The maps correspond to values measured or reconstructed on 6th July 2018.

**Table 2.** Predicted-AOD 0.47µm and 0.55µm obtained from Stage-2 RF models were regressed against measured Satellite-AOD 0.47µm and 0.55µm in a linear regression model. The performance was evaluated using OOB CV together with RMSE (µg/m^3^), intercept (µg/m^3^), and slope (µg/m^3^).

| Stage-2 OOB CV |  |  |  |  |  |  |  |  |
| --- | --- | --- | --- | --- | --- | --- | --- | --- |
| | Predicted-AOD $0.47\mu\text{m}$ | | | | Predicted-AOD $0.55\mu\text{m}$ | | | |
|  | R2 | RMSE | Inter. | Slope | R2 | RMSE | Inter. | Slope |
| 2008 | 0.977 | 0.010 | -0.001 | 1.009 | 0.977 | 0.007 | -0.001 | 1.009 |
| 2009 | 0.976 | 0.010 | -0.001 | 1.010 | 0.976 | 0.007 | -0.001 | 1.010 |
| 2010 | 0.968 | 0.009 | -0.001 | 1.013 | 0.968 | 0.007 | -0.001 | 1.013 |
| 2011 | 0.988 | 0.010 | -0.001 | 1.005 | 0.988 | 0.007 | 0.000 | 1.005 |
| 2012 | 0.980 | 0.010 | -0.001 | 1.008 | 0.981 | 0.007 | -0.001 | 1.008 |
| 2013 | 0.984 | 0.010 | -0.001 | 1.007 | 0.984 | 0.007 | -0.001 | 1.006 |
| 2014 | 0.970 | 0.009 | -0.001 | 1.012 | 0.970 | 0.007 | -0.001 | 1.012 |
| 2015 | 0.972 | 0.009 | -0.001 | 1.011 | 0.973 | 0.007 | -0.001 | 1.011 |
| 2016 | 0.975 | 0.009 | -0.001 | 1.010 | 0.975 | 0.007 | -0.001 | 1.010 |
| 2017 | 0.963 | 0.009 | -0.001 | 1.015 | 0.963 | 0.007 | -0.001 | 1.014 |
| 2018 | 0.969 | 0.010 | -0.001 | 1.013 | 0.969 | 0.007 | -0.001 | 1.013 |
| **Mean** | 0.978 | 0.010 | -0.001 | 1.009 | 0.978 | 0.007 | -0.001 | 1.009 |

### 3.3. Stage-3 results

Stage-3, the main step of the satellite-based machine learning framework, combines the output of Stage-1 and Stage-2 with a list of spatial and spatio-temporal predictors to estimate PM_2.5_ at the locations of the monitors. The relative importance of the predictors for the Stage-3 RF models are ranked in Table 3. The list with the top-15 predictors demonstrates the larger contribution of the more informative spatio-temporal variables (EMEP4UK PM_2.5_, meteorological parameters, BLH, Stage-2 AOD, and NDVI). All the proposed spatially-lagged PM_2.5_ variables were classified as highly important and their ranking positions varied slightly across the displayed years. This suggests the presence of spatial correlations in PM_2.5_ values that are not entirely captured by the other variables.

Table 4 shows the results of the 10-Fold CV Stage-3 RF models by year. The results indicate a good predictive performance of the model throughout the study period. Overall cross-validated R^2^ ranged from 0.704 (2008) to 0.821 (2011), with an average of 0.767. The average prediction error is 4.042 μg/m^3^, with negligible bias in intercept and slope. The inspection of the spatial and temporal contributions shows that the model performs well generally across the two components, displaying a spatial performance drop only for 2008 (0.486) and 2015 (0.579). The cross-validated spatial R^2^ ranges from 0.486 (2008) to 0.746 (2017), while the cross-validated temporal R^2^ from 0.760 (2008) to 0.843 (2011). The two components have an average of 0.658 and 0.795, respectively. The high spatial R^2^ performance across the years demonstrates that the Stage-3 RF models were able to predict the spatial variation of long-term PM_2.5_ across Great Britain with good accuracy. In Appendix B, Table B1 shows the Stage-3 results by season, demonstrating that the seasonal patterns were well described by RF models, although with lower accuracy for the temporal domain in summer, characterized by a lower 10-Fold R^2^. This drop in performance during summer was also seen by other studies [Stafoggia et al., 2019; Stafoggia et al., 2016].

**Table 3.** Relative importance (%) of the predictors in Stage-3 for the first, middle and last years.

| Stage-3 Predictors | 2008 | 2013 | 2018 |
| --- | --- | --- | --- |
| EMEP4UK $PM_{2.5}$ | 32.41 | 32.83 | 36.74 |
| Spatially-lagged hotspot-$PM_{2.5}$ regional | 2.55 | 2.49 | 6.77 |
| Wind direction | 6.35 | 7.33 | 5.34 |
| Spatially-lagged background-$PM_{2.5}$ regional | 1.14 | 6.06 | 4.93 |
| Day of the year | 3.22 | 4.33 | 3.73 |
| Spatially-lagged hotspot-$PM_{2.5}$ local | 3.97 | 1.65 | 3.66 |
| Precipitation | 6.63 | 2.42 | 3.25 |
| BLH 0h | 2.28 | 2.91 | 2.79 |
| Spatially-lagged background-$PM_{2.5}$ local | 0.94 | 3.07 | 2.75 |
| Month | 1.76 | 2.72 | 2.68 |
| 2m Air temperature | 2.60 | 2.93 | 2.65 |
| Wind speed | 3.08 | 3.75 | 2.53 |
| Sea pressure | 3.09 | 2.60 | 2.49 |
| Relative humidity | 1.77 | 1.56 | 1.81 |
| Nearest background monitor distance | 3.10 | 2.30 | 1.78 |

Note: The top 15 predictors in the RF importance ranking order is determined by the year 2018. The importance is measured by the amount of error reduced due to splits of a given predictor over all trees used in the RF ensemble.

**Table 4.**
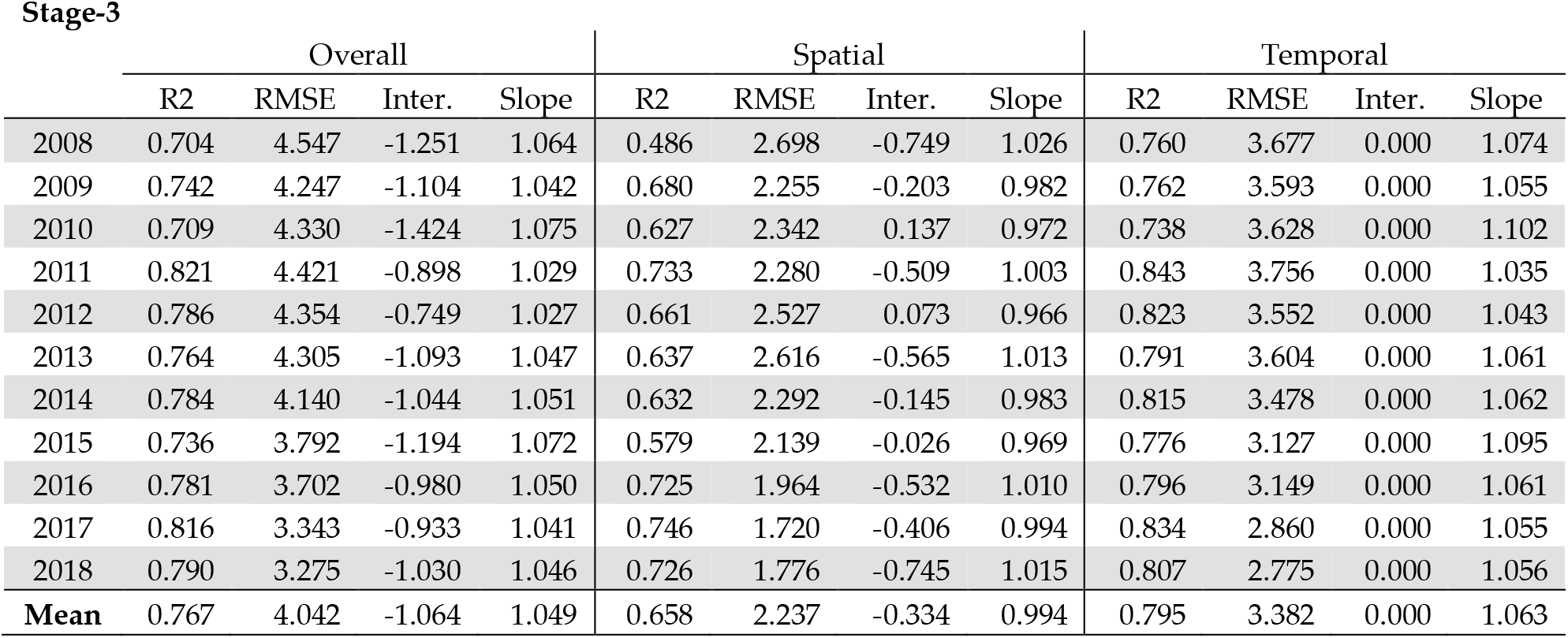
Predicted-PM_2.5_ concentrations obtained from Stage-3 RF models were regressed against Stage-1 measured/predicted-PM_2.5_ concentrations in a linear regression model. The CV-R^2^ (how well the model described the PM_2.5_ variability in new locations) described in three different patterns (overall, spatial, and temporal), RMSE (measure of the model error, µg/m^3^), intercept (µg/m^3^), and slope (µg/m^3^).

| Stage-3 | Overall |  |  |  | Spatial |  |  |  | Temporal |  |  |  |
| --- | --- | --- | --- | --- | --- | --- | --- | --- | --- | --- | --- | --- |
|  | R2 | RMSE | Inter. | Slope | R2 | RMSE | Inter. | Slope | R2 | RMSE | Inter. | Slope |
| 2008 | 0.704 | 4.547 | -1.251 | 1.064 | 0.486 | 2.698 | -0.749 | 1.026 | 0.760 | 3.677 | 0.000 | 1.074 |
| 2009 | 0.742 | 4.247 | -1.104 | 1.042 | 0.680 | 2.255 | -0.203 | 0.982 | 0.762 | 3.593 | 0.000 | 1.055 |
| 2010 | 0.709 | 4.330 | -1.424 | 1.075 | 0.627 | 2.342 | 0.137 | 0.972 | 0.738 | 3.628 | 0.000 | 1.102 |
| 2011 | 0.821 | 4.421 | -0.898 | 1.029 | 0.733 | 2.280 | -0.509 | 1.003 | 0.843 | 3.756 | 0.000 | 1.035 |
| 2012 | 0.786 | 4.354 | -0.749 | 1.027 | 0.661 | 2.527 | 0.073 | 0.966 | 0.823 | 3.552 | 0.000 | 1.043 |
| 2013 | 0.764 | 4.305 | -1.093 | 1.047 | 0.637 | 2.616 | -0.565 | 1.013 | 0.791 | 3.604 | 0.000 | 1.061 |
| 2014 | 0.784 | 4.140 | -1.044 | 1.051 | 0.632 | 2.292 | -0.145 | 0.983 | 0.815 | 3.478 | 0.000 | 1.062 |
| 2015 | 0.736 | 3.792 | -1.194 | 1.072 | 0.579 | 2.139 | -0.026 | 0.969 | 0.776 | 3.127 | 0.000 | 1.095 |
| 2016 | 0.781 | 3.702 | -0.980 | 1.050 | 0.725 | 1.964 | -0.532 | 1.010 | 0.796 | 3.149 | 0.000 | 1.061 |
| 2017 | 0.816 | 3.343 | -0.933 | 1.041 | 0.746 | 1.720 | -0.406 | 0.994 | 0.834 | 2.860 | 0.000 | 1.055 |
| 2018 | 0.790 | 3.275 | -1.030 | 1.046 | 0.726 | 1.776 | -0.745 | 1.015 | 0.807 | 2.775 | 0.000 | 1.056 |
| Mean | 0.767 | 4.042 | -1.064 | 1.049 | 0.658 | 2.237 | -0.334 | 0.994 | 0.795 | 3.382 | 0.000 | 1.063 |

### 3.4. Stage-4 results

Stage-4 provides the prediction of daily PM_2.5_ concentrations for each of the 234.429 1 km grid covering Great Britain. Results indicate an annual PM_2.5_ average of 9.41 μg/m^3^ for 2008, 10.17 μg/m^3^ for 2013, 8.05 μg/m^3^ for 2018, and 8.84 μg/m^3^ for 2008-2018 but with a strong spatial and temporal variation. The spatial distribution of annual average PM_2.5_ concentrations for 2008, 2013, and 2018 are shown in Figure 3, revealing a decrease of pollution levels in recent years across the whole territory, although slightly stronger in England. Table C1 in Appendix C provides the same figures for all the years, confirming the decreasing trend. The spatial comparison suggests that PM_2.5_ concentrations are lower in Scotland and Wales compared to the more populated southern regions of England, with hotspots located in urban areas such as Liverpool, Manchester, Birmingham, and Greater London. At the bottom of Figure 3, the maps display the corresponding annual average of PM_2.5_ levels in London, demonstrating the precision of the multi-stage ML model in reconstructing PM_2.5_ concentrations in 1 km grid cells within urban areas. The maps show local hotspots of high pollution, with a spatial distribution that however changes along the study period.

**Figure 3:**
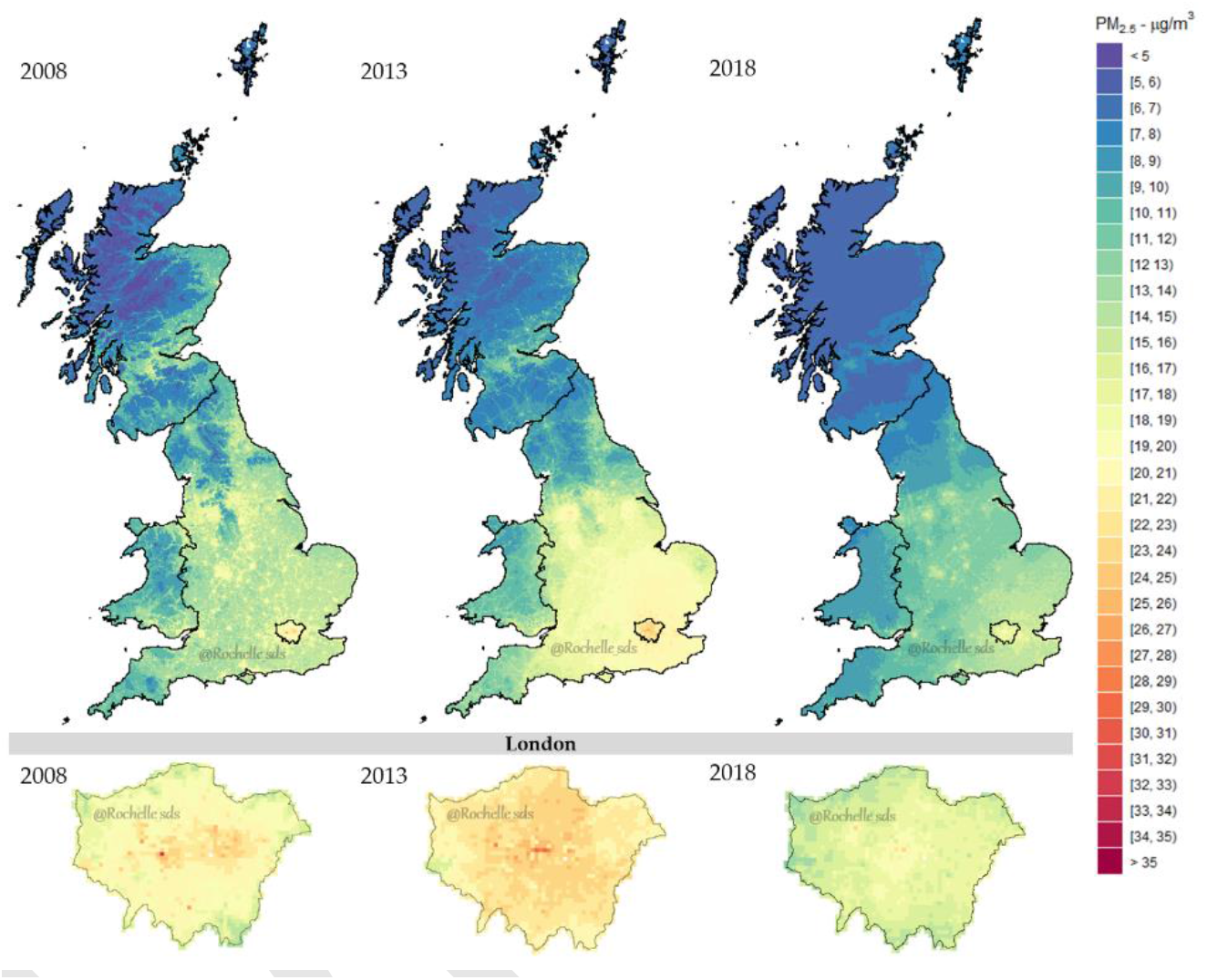
Stage-4 predicted PM_2.5_ concentrations across Great Britain (Top) and London (Bottom) for 2008, 2013, and 2018 aggregated by annual means. All plots were built under the same colour scale.

The greatest contribution of this study is the ability of the satellite-based ML models to reconstruct daily levels of PM_2.5_ over a 1 km grid across a wide geographical domain. Figure 4 displays the spatial distribution of PM_2.5_ estimations across Great Britain (Top) and London (Bottom) for specific days within the study period. It is interesting to note the wide variation in PM_2.5_ concentrations between days in the same area and between areas in different days. For instance, the maps in the left panels represent a day with almost no variation and generally low concentrations; the maps in the mid panels display a strong north/south split likely linked to weather conditions; the right panels show a more complex pattern with wider range of PM_2.5_ values, a large area with very high pollution concentrations located in east London, and hotspots in highly-populated urban areas across England.

**Figure 4:**
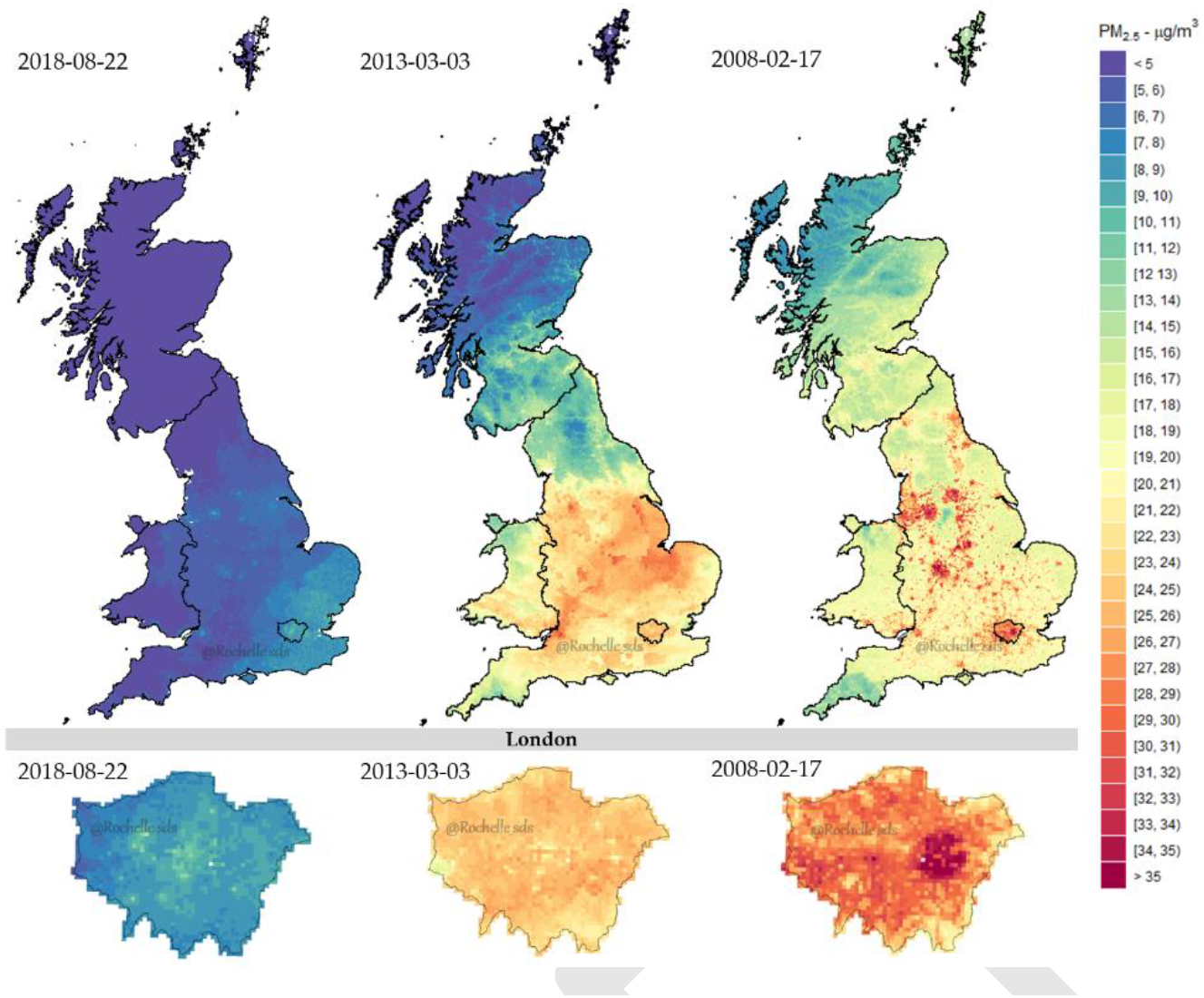
Stage-4 day-specific PM_2.5_ estimations across Great Britain (Top) and London (Bottom).

## 4. Discussion

This study presents the first application of satellite-based spatio-temporal ML methods to reconstruct levels of pollution across Great Britain, providing estimates of daily PM_2.5_ concentrations over a 1 km grid during 2008-2018. The multi-stage ML framework provided significant advantages, allowing the combination of information from multiple data sources, such as air quality monitoring networks, remote sensing satellite products, chemical dispersion models, reanalysis databases, administrative census data, among others.

The beginning of the study period (2008 and 2009) had lower quantity of monitors measuring PM_2.5_ across Great Britain, but this number increased considerably from 2010 after a new measuring network has been established in 2009 [DEFRA, 2012]. Therefore, Stage-1 was an extremely important step to extend the number of PM_2.5_ measurements. The implementation of Stage-2 was also relevant to fill the gaps in MAIAC AOD retrievals from satellites, thus maximizing the available information. Stage-3 produced a ML prediction model based on a long list of informative predictors, accounting for potentially complex inter-relationships and functional forms. The Stage-3 ML algorithms offered an excellent performance, showing an average cross-validated R^2^ of 0.767 across the period, with increased predictive ability in the last years. Stage-4 provided a single PM_2.5_ estimation for each of the 234.429 1 km grid cells in each of 4,018 days, totaling around 950 million data points. The methodology provides a complete spatial coverage, high resolution, and relative small error of the predictions, coupled with and the ability to capture variations in PM_2.5_ concentrations across both spatial and temporal domains. The model offers a prediction accuracy that makes the output suitable for application in epidemiological studies on short and long-term health effects of air pollution.

The output from the empirical ML model developed in this study complements existing databases of modelled PM_2.5_ in the United Kingdom, with some advantages for applications in epidemiological studies. Country-wide maps generated by emission-dispersion models are usually available at coarser spatial or temporal resolution [DEFRA, 2020; EMEP4UK, 2020; Savage et al., 2013, and generally they show lower small-scale accuracy when tested against observed monitoring data [Hood et al., 2018; Lin et al., 2017]. The spatio-temporal ML models presented here demonstrated comparable predictive performance to similar methods applied in other countries, based either on single-learner ML models [de Hoogh et al., 2018; Stafoggia et al., 2019], ensemble ML models [Chen et al., 2018; Di et al., 2019; Yazdi et al., 2020], or generalised additive models (GAM) [Kloog et al., 2015]. Yazdi et al. (2020) developed an ensemble ML model (composed by RF, Deep Neural Network, GAM, Gradient Boosting, K-nearest Neighbour) to estimate PM_2.5_ for Greater London, reaching mean 2005-2013 CV spatial-R^2^ of 0.396. Using 2008-2013 period, this study reached CV spatial-R^2^ of 0.637 for the whole Great Britain. Modelling a larger area might have provided more information and higher spatial variability, improving substantially the CV spatial-R^2^. The application of a single learner to model air pollution in both spatial and temporal domains for Great Britain achieved a satisfactory performance. Nonetheless, ensemble model formats can be used in future applications using the same area of study and variables to assess how much performance gain is reached compared to the RF-only learner.

In 2008, a new Air Quality Directive (AQD) came into force bringing considerable changes to the following UK annual air quality assessment in 2010, setting an annual mean target of 25 µg/m^3^ [Brookes et al., 2011]. Several policy controls and emission reductions had been put in place aiming to reduce from 2010 the road traffic PM emission by 83%, off-road mobile machinery by 54%, and energy production by 32% until 2020 [AQEG, 2013]. Independently of the temporal aggregation (daily or annual), all maps shown in this study detected this considerable drop in the PM_2.5_ concentrations from 2010 across Great Britain.

Some limitations must be acknowledged. First, the multi-stage model relies on the extension of the observed series of PM_2.5_ by predicting values based on co-located PM_10_ in Stage-1. This step was necessary for the application of the method in early years characterized by sparse PM_2.5_ monitoring, which is likely to have contributed to the lower predictive performance in this period. In addition, the cross-validation procedures revealed the presence of some bias in the Stage-1 predictions, particularly in the spatial domain, probably linked to limitations in modelling the relative distribution of the two PM components. Second, while the model displayed a good performance throughout the period, the accuracy is worse in the temporal domain in summer (Table B1), suggesting limitations in capturing the higher temporal variation in this season. Third, the generalization of the prediction model was dependent on the selected locations of the monitors, which may be not representative of the study domain. This can result in an underestimation of the error and potentially biases in the predictions in more remote and less represented areas, if structural spatial differences are not entirely captured by the model covariates.

Future research directions for improving the current definition of the spatio-temporal ML model and its output can be discussed as well. First, availability of predictors that capture spatio-temporal variations is critical for ensuring high predictive ability. Improvements in emission-dispersion models such as EMEP4UK, whose output was consistently selected as the most important predictor, or in remote sensing satellite products, which in contrast show limited contributions at present, can be key factors for increasing the model performance. For instance, new instruments available in more recent earth observation satellites (e.g., Copernicus Sentinel-5 Precursor) are explicitly developed for applications in pollution measurements and environmental health, and can be integrated in future versions of the model [ESA, 2020]. Second, spatially-lagged PM_2.5_ variables, computed using IDW-LOOCV, were selected within the top ten-best predictors, suggesting that the performance can be extended further if the underlying spatial correlation structure is appropriately modelled. This can be achieved by extending the current spatial definition to fully spatio-temporal variables, and by applying more sophisticated spatial methods to incorporate such correlation. Third, published articles have presented alternative ML algorithms in single-learner [Just et al., 2018; Zan et al., 2017] or ensemble models [Di et al., 2019], which can be tested and applied in alternative to the RF method proposed here. Fourth, finally, previous studies have developed methods to increase the resolution of the predictions beyond the 1 km grid, for instance using additional stages involving small-area variables [de Hoogh et al., 2018; Di et al., 2019; Stafoggia et al., 2019]. Similar approached can be applied here.

## 5. Conclusions

This study developed and applied a multi-stage ML model, combining data for multiple sources, including remote sensing satellite products, climate and atmospheric reanalysis models, chemical transport models, and geospatial features, to generate a complete map of daily PM_2.5_ concentrations in 1 km grid across Great Britain in the period 2008-2018. The model showed good performance overall and in both spatial and temporal domains, with an accuracy that is compatible with the use of such reconstructed values as proxy for PM_2.5_ exposures in epidemiological studies. In particular, the availability of high-resolution measures that can be linked as such or aggregated at different spatial and/or temporal scales makes the output suitable for investigations on both transient and chronic health risks associated to short and long-term exposures to PM_2.5_, respectively.

### Funding

This study was supported by the Medical Research Council-UK (Grant ID: MR/M022625/1), the Natural Environment Research Council UK (Grant ID: NE/R009384/1), and the European Union’s Horizon 2020 Project Exhaustion (Grant ID: 820655). EMEP4UK Model results and contributions by S.R. and M.V. were supported by award number NE/R016429/1 as part of the UK-SCAPE programme delivering National Capability.

## Data Availability

All data used to perform the analysis are in the public domain. References and sources are provided in the text.

## Acknowledgments

The authors are grateful for the technical support received from the European Centre for Medium-Range Weather Forecasts (ECMWF). The authors also would like to thank the European Space Research Institute (ESRIN) from the European Space Agency (ESA) for their feedback on this study. This research had free access to all data sources: (i) NASA EOSDIS Land Processes Distributed Active Archive Center (LP DAAC), (ii) NOAA National Centers for Environmental Information (NCEI), (iii) Copernicus Land Monitoring Service (CLMS), (iv) Copernicus Atmosphere Data Store, (v) Copernicus Climate Data Store, (vi) EMEP4UK, (vii) UK Department for Environment, Food & Rural Affairs.

